# Applying Topical Anesthetic on Pediatric Lacerations in the Emergency Department: A Quality Improvement Project

**DOI:** 10.1101/2021.07.06.21260013

**Authors:** Nagham Faris, Mohamad Mesto, Sandra Mrad, Ola El Kebbi, Noor Asi, Rasha D. Sawaya

## Abstract

**Background:** Caring for pediatric lacerations in the Emergency Department (ED) is typically painful because of irrigation and suturing. To improve this painful experience, we aimed to increase the use of a topical anesthetic, Eutectic Mixture of Local Anesthetics (EMLA) on eligible pediatric lacerations with an attainable, sustainable, and measurable goal of 60%.

**Local Problem:** The baseline rate of applying topical anesthetic to eligible lacerations was 23% in our ED. We aimed to increase the use of topical anesthetics on eligible pediatric lacerations to a measurable goal of 60% within 3 months of implementing our intervention.

**Methods:** We conducted a prospective, single center, interrupted time series, ED quality improvement project from November 2019 to July 2020. A multidisciplinary team of physicians and nurses performed a cause-and-effect analysis identifying two key drivers: early placement of EMLA and physician buy-in on which we built our Plan, Do, Study, and Act (PDSA) cycles. We collected data on number of eligible patients receiving EMLA, as well as patient and physician feedback via phone calls within 2 days post encounter. Balancing measures included ED length of stay (LOS), patient and physician satisfaction with EMLA, and side effects of EMLA.

**Results:** We needed 3 PDSA cycles to reach our goal of 60% in 3 months, which was also maintained for 5 months. PDSA cycles used educational interventions, direct provider feedback about non-compliance and patient satisfaction results obtained via phone calls. Balancing measures were minimally impacted: 75% good patient satisfaction, No adverse events but an increase in LOS of patients who received EMLA compared to those who did not (1.79 ± 0.66 VS 1.41 ± 0.83 hours, p<0.001). The main reasons for dissatisfaction for physicians were the increased LOS and the preference for procedural sedation or intranasal medications.

**Conclusion:** With a few simple interventions, our aim of applying EMLA to 60% of eligible pediatric lacerations was attained and maintained.

## Introduction

Pediatric traumatic lacerations are a common Emergency Department (ED) chief complaint; many requiring suturing. This painful procedure can be alleviated with a topical anesthetic or an injection of a local anesthetic, which is however also in itself painful. Topical anesthetics alone may offer similar analgesia to that of local needle anesthetic infiltration especially for lacerations of the face and scalp. Moreover, topical anesthetics will decrease the pain of injection of local anesthetics in conditions where topical anesthetics alone are not enough for analgesia, such as deeper wounds [1-3]. Topical anesthetics can also be helpful prior to the use of tissue adhesives preventing discomfort during their application and making wound irrigation more tolerable [4]. The American Academy of Dermatology 2016 guidelines recommend the use of topical anesthetics as first-line anesthetics for laceration repairs in children [1]. Moreover, applying topical anesthetics to wounds requiring suturing or stapling is a quality measure used to assess performance of pediatric EDs [5].

There are multiple topical anesthetic creams including Eutectic Mixture of Local Anesthetics (EMLA), Lidocaine-Epinephrine-Tetracaine (LET) and Tetracaine, Adrenaline, and Cocaine (TAC). Although the use of EMLA on non-intact skin is off label, it has been extensively studied over the past 2 decades showing significant benefit from the application of EMLA prior to laceration repair without any increase in harm compared to other topical anesthetics [3, 6-10]. A randomized, double-blind, controlled study comparing the use of EMLA and LET on simple lacerations showed no significant difference in pain due to the subsequent infiltration of local lidocaine [7]. Zempsky et al compared the use of EMLA and TAC on extremity lacerations in children between 5 and 18 years of age and found that wounds to which EMLA was applied had better anesthesia effect and required less supplemental anesthetics [11]. Note that the use of TAC has decreased because of toxicity related to cocaine [3].

No significant increase in risk of systemic side effects or complications were reported due to the application of EMLA on non-intact skin [2, 6, 11]. Local temporary side effects might include contact dermatitis, erythema or pallor, irritation of the eyes, and edema [3, 12]. These are the same side effects as those noted on intact skin. Methemoglobinemia is a serious potential side effect of topical anesthetics on intact or non-intact skin, but the risk is low and was reported to be 0.035% in a study by Chowdhary et al [13]. Risk of methemoglobinemia is slightly higher in infants below 3 months of age and in patients receiving other drugs that form methemoglobin such as paracetamol, nitrous oxide, sulfonamides, phenobarbital, and phenytoin [3, 12].

### Objectives

We implemented a Quality Improvement Project aiming to apply a topical anesthetic cream (EMLA) on the lacerations of pediatric patients in order to improve our pediatric patients’ experience during laceration repair, in alignment with our over-arching goal to create a pediatric friendly, pain-free environment in our ED.

Two months prior to our intervention start date, we collected data on the use of EMLA on all lacerations in patients 19 years old or less. The baseline rate of applying topical anesthetic to eligible lacerations was 23%. We aimed to increase the use of topical anesthetics on eligible pediatric lacerations (3 months to 19 years old) to an attainable, sustainable, and measurable goal of 60% within 3 months of implementing our intervention.

## Methods

### Design and setting

This was a prospective, single center, interrupted time series design, quality improvement project aimed at increasing the use of topical anesthetics for pediatric lacerations presenting to the ED, using Plan, Do, Study, Act (PDSA) cycles, from November 20, 2019 until July 20, 2020. It was exempt from the American University of Beirut Institutional Review Board and approved by the American University of Beirut Medical Center (AUBMC) Quality and Safety committee.

The project took place in the ED at the AUBMC, a tertiary care hospital in Lebanon, pre-COVID-19 pandemic. AUBMC is a 350-bed hospital, with the largest ED in the country. The ED at AUBMC consists of 43 beds and is divided into three sections: high acuity adults, low acuity adults, and pediatrics, each with dedicated teams. We care for approximately 57000 ED patients/year, 25% of which are less than 19 years of age.

Eligible patients included all children with a laceration that fit the following criteria: children 3 months to 19 years of age [3, 12], simple, uncomplicated, linear and clean lacerations [4, 6, 7] that are less than 5 cm long [11]. We excluded patients that had wounds involving mucous membranes [4], that required compression, deep wounds with obvious injury to tendons, cartilage, bones, or vessels [4], abrasions or superficial wounds that are unlikely to require repair [4], allergies to EMLA or any of its components [4, 6, 7, 11], intake of medications that might predispose to methemoglobinemia (paracetamol, nitrous oxide, sulfonamides, phenobarbital, and phenytoin) [3, 7, 11, 12], application or infiltration of an anesthetic prior to ED presentation [4], polytrauma [6], animal or human bites [6, 7, 11], puncture wounds [7], devitalized tissue [6, 7], infected or heavily contaminated lacerations [6, 7], lacerations more than 6 hours old [4, 6, 7] or a rash [6, 7].

### SMART aim

The aim of this project was to reach the goal of applying topical anesthetics to at least 60% of the eligible patients described above, presenting to the ED, within 2 months of implementing our intervention, and to maintain this goal for at least 5 months thereafter.

In the pediatric EDs of the United States, the percentage of pediatric patients with head, face, and neck lacerations who receive a topical anesthetic prior to wound repair is around 40% [5]. Given that there are no published standards for the use of topical anesthetics for lacerations and that this is not a life-saving intervention, we chose 60% as an achievable and sustainable goal. Specifically, the outcome measure was the percentage of patients satisfying inclusion criteria who got EMLA prior to repair (goal of 60%).

Data was obtained from a report generated on the electronic medical record. To calculate the percentage of patients receiving EMLA, the numerator was the number of patients who had a Registered Nurse (RN) or Medical Doctor (MD) medication order for EMLA, and the denominator was the number of patients with lacerations satisfying the inclusion criteria, based on chart reviews.

Balancing measures were ED length of stay (LOS), patient and physician satisfaction with EMLA, and side effects of EMLA. Given that EMLA’s time to peak effect is 45 minutes to 1 hour, we aimed to measure its impact on the ED LOS. The information regarding patient and physician satisfaction as well as any possible side effects was obtained through phone calls to the family and physician who sutured the patient within 48 hours of discharge.

### Intervention

Using the Institute for Health Improvement’s (IHI) Model for Improvement guidelines [14], we assembled a multidisciplinary team (EMLA team) of ED MDs and RNs to review causes behind the low use of topical anesthetics for lacerations and obstacles that may be faced to reach our objective.

We compared the availability and cost of different topical anesthetics. Obtaining the ingredients for our pharmacy to mix LET was too expensive for our ED and patient population, therefore we opted for EMLA, albeit the longer time to onset of action, which is around 45 minutes to 1 hour [4, 11].

The EMLA team findings were interred into a cause-and-effect analysis diagram (Figure 1), and two key drivers were identified: placing the topical anesthetic cream as soon as possible and MD (including non-ED physicians who are consulted e.g plastic surgeons) buy-in.

**Figure 1:**
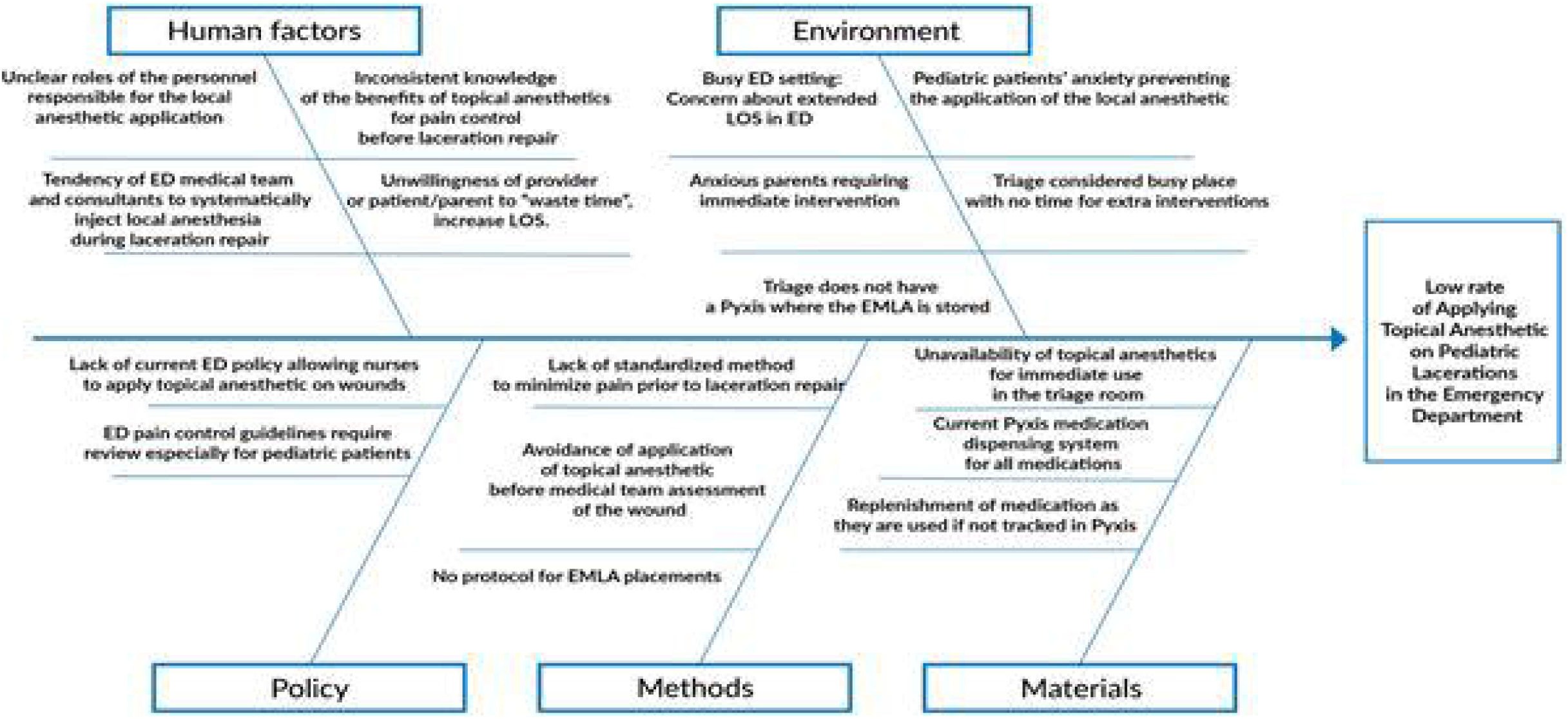
Cause-and-Effect Diagram. LOS: Length of stay, ED: Emergency Department, EMLA: Eutectic Mixture of Local Anesthetics Pyxis: automated medication dispensing system

**Figure 2:**
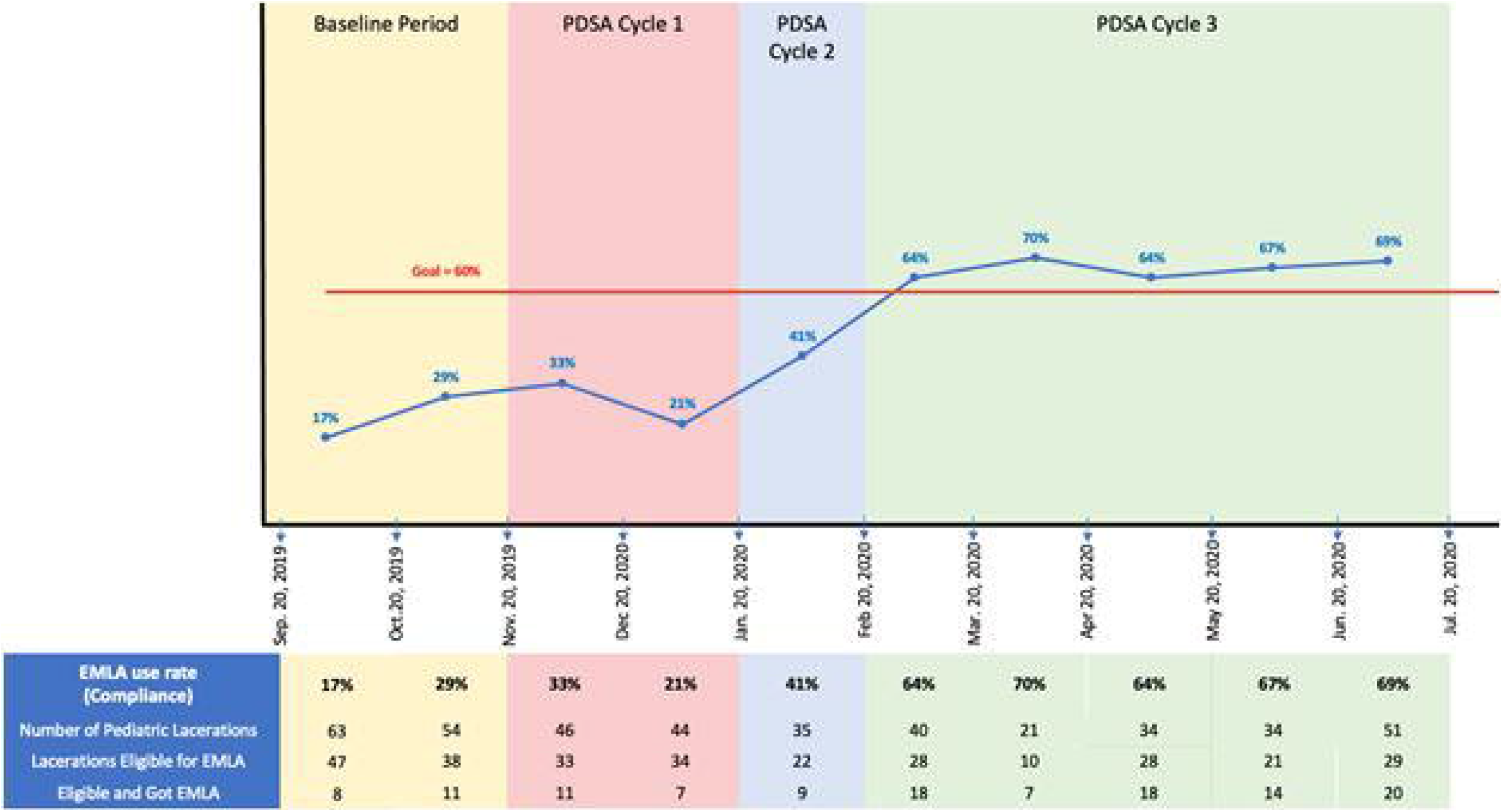
Timeline of interventions and the effect on main outcome.

In our first key driver, we hypothesized that we could increase EMLA use by placing it at as early as possible to counter act its long duration of action. After discussions with key players, we noted that we could not place EMLA in triage, but that the modular RN (RN caring for the patient in the assigned treatment room) could. Therefore, the EMLA team developed a policy on September 7, 2019, where RNs can order and place the topical anesthetic cream if inclusion criteria are satisfied. The workflow included the RNs deciding on the need for EMLA and placing it to eligible lacerations soon after the patient’s presentation to the ED. A poster for inclusion/exclusion criteria was placed in the triage area and in the pediatrics section of the ED and all RNs were educated about the project and their role. The policy was then implemented.

In our second key driver, we hypothesized that getting MD buy-in will increase the use of EMLA. Our 1st key driver was to get the modular RNs to place the EMLA; however, sometimes, in the busy shifts, the MD may see the patient before the modular RN can assess him/her. Therefore, the MDs were also educated using posters, emails, and a presentation during resident and attending meetings about this project. Physicians e.g plastic surgeons who may be consulted to repair the lacerations were also informed and educated of this change in process, to increase acceptance. The outcome measure for these 2 key drivers was the frequency of EMLA ordered.

### Data Collection

We identified patients weekly, through an electronic report including any patient who had topical skin adhesive (Dermabond), thin adhesive bandages, or sutures charged. The charts were reviewed for inclusion criteria and the following data was collected: order of EMLA, LOS, basic patient demographics, and laceration repair done by a consultant (plastic or otolaryngology team).

For a period of 3 months during the implementation phase of this project, between January 2020 and March 2020, we called back, within 48 hours, the ED provider and legal guardian of every patient on whom EMLA was used to determine their satisfaction with the process, and to identify unforeseen problems or side effects from the EMLA application.

### Data Analysis

Descriptive statistics were summarized by presenting the number and percentage for categorical variables and mean and standard deviations for continuous variables. The association between EMLA and other categorical variables was carried out by using the Fisher’s exact test. Independent t-test was used for the association with continuous variables. P-value <0.05 was used to indicate statistical significance. All statistical analyses were performed using the Statistical Package for Social Sciences (SPSS, version 24).

## Results

From September 20, 2019, until July 20, 2020, 422 patients with age 3 months to 19 years presented to our ED with lacerations. Among those, 290 lacerations were eligible for EMLA application. **Error! Reference source not found**. shows the characteristics of patients presenting with lacerations eligible for EMLA. Most eligible lacerations were on the face (232/290, 80%). Among eligible lacerations, 43 required a specialist consultation for laceration repair (such as plastic surgery or otolaryngology). Patients who got EMLA were significantly younger (5.03 ± 3.49 VS 7.78 ± 5.63 years, p<0.001) and had increased LOS (1.79 ± 0.66 VS 1.41± 0.83 hours, p<0.001).

### PDSA Cycle 1

As of November 20, 2019, the QI project was introduced to the ED team in meetings with residents, attendings, and nurses. Posters delineating the eligibility criteria and application instructions as well as ones encouraging patients to ask for EMLA, were placed in the pediatric section of the ED. This educational intervention did not increase our EMLA placement significantly (see **Error! Reference source not found**.). We noticed resistance from certain physicians and hesitancy from nurses.

### PDSA Cycle 2

On January 20, 2020 we started a chart review of patients with lacerations within 48 hours of discharge. For eligible lacerations, we called the physician who performed the repair to obtain feedback when EMLA was used and if not used, the reasons why. We reemphasized the benefit of EMLA and the goal of a pain-free pediatric ED. We called the patients’ parents who received the topical anesthetic to ask about satisfaction and any adverse effects. Data from this intervention was shared with the clinicians. This intervention increased our application rate to 41% (see **Error! Reference source not found**.).

### PDSA Cycle 3

On February 9, 2020, we started calling the family of patients who had eligible lacerations but did not receive EMLA, to know if they would have liked receiving it. This was used to provide feedback to the caring physicians. On February 12, 2020, reminders were sent to residents as different residents were now working in the ED. Through the chart review, we identified the RNs who did not order EMLA and had one-on-one discussions with them about barriers, reviewed our policy, and empowered them. Forgetting the EMLA was a common concern, so desktop wallpapers and more posters were added to the pediatric section of the ED. This increased our numbers to > 60% (see **Error! Reference source not found**.).

### Main Outcome

The goal of applying EMLA to 60% of eligible pediatric lacerations was achieved within 3 months and sustained for 5 months.

### Balancing Measures

Patients who got EMLA had a significantly higher mean LOS than those who did not (1.81 hours ± 0.731 and 1.41 hours ± 0.845 respectively, p<0.0001) as shown in **Error! Reference source not found**.. Yet, the majority (18/24, 75%) of the patients (or parents) interviewed reported a good overall experience with the application of topical anesthetics and did not complain regarding the increased LOS. In fact, 80% of patients who did not get EMLA, would have wanted it. No adverse effects were reported (See **Error! Reference source not found**.).

Physicians (n=35) showed a variable degree of satisfaction with EMLA. 19/35 physicians (54%) were satisfied and 16/35 (46%) were not. Reasons for their dissatisfaction included: EMLA did not help relieve patients’ anxiety, the use of EMLA required a lot of time, and physician’s preference for procedural sedation to relieve the child’s pain and anxiety.

## Discussion

In the pediatric ED, simple but painful procedures, such as laceration repairs, are frequently performed. To improve patients’ experience when undergoing laceration repairs, we implemented this ED quality improvement project aiming to apply the safe and effective topical anesthetic (EMLA) to eligible pediatric lacerations.

Through this quality improvement project, by working in a multidisciplinary team, developing a cause-effect diagram we successfully used three PDSA cycles to achieve and sustain our goal of applying EMLA to at least 60% of the eligible pediatric patients with lacerations, with minimal impact to balancing measures except for a significant increase in the patient’s length of stay but still with very good patient satisfaction with the process, and safety profile.

This manuscript is significant, since, to our knowledge, little has been documented regarding QI projects on the use of topical anesthetics, specifically EMLA. Sherman et al performed a QI project aiming to apply LET to pediatric facial lacerations by implementing educational initiatives. Their interventions that included educational sessions and posters increased the rate of application LET and reduced the time to its application[15]. In our project, we show that with a few simple PDSA cycles, the ED team can safely introduce a new process that would decrease pediatric pain in the ED. Similar to Sherman et al, we used educational initiatives to increase the use of EMLA; however, time of application was not assessed since the use of topical anesthetics on non-intact skin was not common practice in our setting before the introduction of our project. We also used the positive feedback of patients’ families to encourage the use of this topical anesthetic.

Having a multidisciplinary team including RNs and MDs was key for the success of this project. All RNs and MDs were aware of the project soon after its introduction. Since EMLA needs at least 45 minutes for onset of action, MDs and RNs were both working together to apply EMLA as soon as possible. In a similar QI project done in New York and aiming to apply LET to pediatric facial lacerations, both RNs and MDs received education about the project in multidisciplinary meetings, and both aimed to apply LET the soonest, depending on their volume and staffing. If LET was not placed in triage, the MD or another RN would place it inside the ED[15]. In our ED, we were not able to place EMLA in triage, and therefore, the RN would place it on the laceration shortly after the patient enters the ED. However, MDs were helping to place it soon if the RN was busy. Cregin et al developed a topical analgesia protocol to decrease pain in nonurgent painful procedures for pediatric patients. A multidisciplinary approach involving leadership, MDs, and RNs increased compliance with the newly developed protocol[16].

Physician buy-in was a barrier in our project. Some physicians believed that EMLA did not relieve patients’ anxiety, is not worth the wait, and that sedation can be used instead. In order to overcome this barrier and to improve physician buy-in, the team communicated directly to the physicians and explained the importance of this project by highlighting the good patient satisfaction, the absence of side effects, and the willingness of patients and families to wait for EMLA’s onset of action. Those were strong arguments to encourage physicians to use this topical anesthetic. Similarly, in a community medical center in New York, after the development of an analgesia protocol for nonurgent pediatric procedures, MDs were not fully compliant because they did not wait for the analgesia effect. Leaders within departments had one-on-one communications with noncompliant physicians to reeducate about the importance of the intervention. This improved physicians’ buy-in and compliance [16].

Moreover, although procedural sedation in the ED is generally safe and gives better anxiety and pain relief, side effects sometimes occur and include hypoxia, apnea, and vomiting [17]. Also, patients receiving procedural sedation usually have a longer LOS and higher costs [18]. For safe and effective anxiolysis during laceration repair, intranasal midazolam can be used [19] in addition to the topical anesthetic which provides analgesia.

The time needed for adequate anesthesia after the application of EMLA and before wound repair is around 1 hour. Of note, a study done in an ED in Korea showed that patient LOS did not increase significantly with pretreatment of lacerations with EMLA as patients were waiting in any case to see the doctor [6]. In our patient population, the LOS of those who got EMLA was significantly higher than that of patients who did not. This may be because our time to doctor is in general low. For pediatric patients, the average door to physician waiting time in our ED is 10 minutes (administrative data). Although statistically significant, the difference in mean LOS was only 0.38 hours. Based on our call-backs, the increased LOS was not perceived negatively by families. Also, we believe that the benefits of this intervention and the positive feedback of families are worth this slight increase. Therefore, it is important to develop protocols and apply EMLA in triage as soon as patients arrive [4, 11] especially in our setting where the waiting time to see the physician in the ED is very short.

There are some limitations to our study. First, data was obtained through retrospective chart reviews, and therefore, might be incomplete or inaccurate. However, the data collected was mostly objective data, not requiring any interpretation. The subjective data was collected prospectively via phone calls. Second, this project was a single center project, implemented in a hospital in Beirut, Lebanon. Our results might not be generalizable to other patient populations and EDs Nevertheless, the barriers faced and lessons learnt are useful to all settings wishing to implement such a QI project.

In conclusion, with few simple interventions, we were able to reach and sustain our goal of placement of EMLA in 60% of eligible patients helping us create a pain-free ED. Physician buy-in and the duration of action of EMLA were the main barriers. However, patient satisfaction with the outcome is a metric worth pursuing. The choice of anesthetic and method of implementation will depend on the setting. The lessons learnt can be applied to other projects and settings. Educational-based initiatives are helpful to introduce unfamiliar treatments or processes and encourage their use. Also, the collaboration of nurses and physicians in multidisciplinary approach can improve compliance and help reach the goal.

**Table 1:** Characteristics of pediatric patients presenting with eligible lacerations within the study period.

| Characteristics of Patients |  | EMLA<br>n (%) | NO EMLA<br>n (%) | p-value |
| --- | --- | --- | --- | --- |
| Age (years) mean ± SD | | 5.03 ± 3.49 | 7.78 ± 5.63 | <0.001 |
| Gender | Male | 76 (40) | 114 (60) | 0.252 |
|  | Female | 47 (47) | 53 (53) |  |
| Laceration Location | Face | 103 (44.4) | 129 (55.6) | 0.172 |
|  | Extremity | 20 (34.5) | 38 (65.5) |  |
| Specialty Consults for Laceration Repair |  | 36 (83.7) | 7 (16.3) | 0.009 |
| Length of Stay (hours) mean ± SD | | 1.79 ± 0.66 | 1.41 ± 0.83 | <0.001 |

SD : standard deviation

**Table 1:**
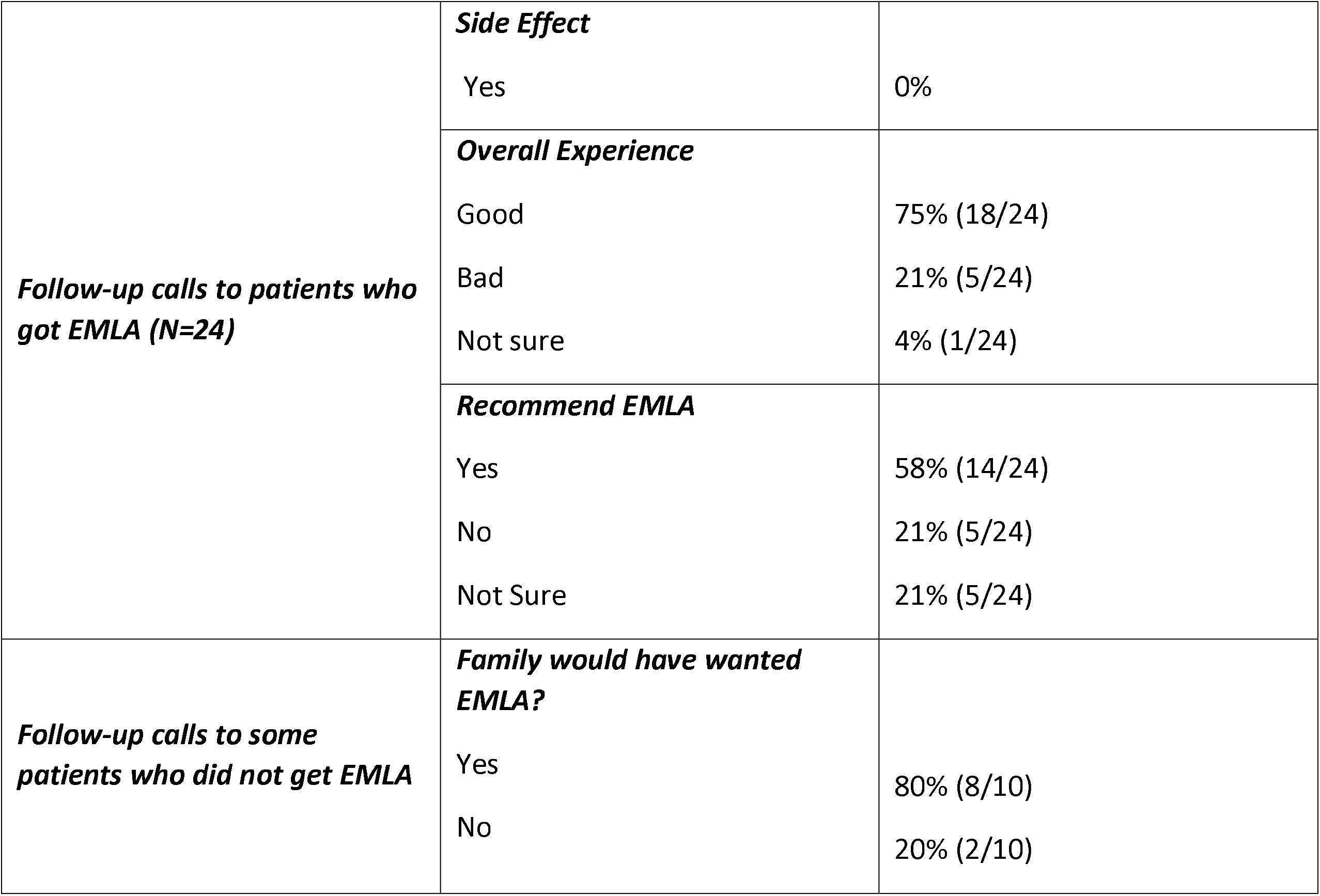
Results of follow-up calls to patients.

## Data Availability

Data was extracted from electronic patient charts.

## References

1. Kouba, D.J., et al., Guidelines for the use of local anesthesia in office-based dermatologic surgery. J Am Acad Dermatol, 2016. 74(6): p. 1201–19.

2. Ferguson, C., B. Loryman, and R. Body, Best evidence topic report. Topical anaesthetic versus lidocaine infiltration to allow closure of skin wounds in children. Emerg Med J, 2005. 22(7): p. 507–509.

3. Young, K.D., Topical anaesthetics: What’s new? Archives of disease in childhood - Education & practice edition, 2015. 100(2): p. 105–110.

4. Priestley, S., et al., Application of topical local anesthetic at triage reduces treatment time for children with lacerations: a randomized controlled trial. Ann Emerg Med, 2003. 42(1): p. 34–40.

5. Michelson, K.A., et al., Use of a National Database to Assess Pediatric Emergency Care Across United States Emergency Departments. Acad Emerg Med, 2018. 25(12): p. 1355–1364.

6. Park, S.W., et al., Topical EMLA Cream as a Pretreatment for Facial Lacerations. Archives of Plastic Surgery, 2015. 42(1).

7. Singer, A. and M.J. Stark, LET versus EMLA for Pretreating Lacerations: A Randomized Trial. Acad Emerg Med, 2001. 8(3): p. 223–230.

8. Eidelman, A., et al., Comparative efficacy and costs of various topical anesthetics for repair of dermal lacerations: a systematic review of randomized, controlled trials. J Clin Anesth, 2005. 17(2): p. 106–116.

9. Mankowitz, S.L., Laceration Management. J Emerg Med, 2017. 53(3): p. 369–382.

10. Kundu, S. and S. Achar, Principles of Office Anesthesia: Part II. Topical Anesthesia. Am Fam Physician, 2002. 66(1): p. 99–102.

11. Zempsky, W. and R. Karasic, EMLA Versus TAC for Topical Anesthesia of Extremity Wounds in Children. Ann Emerg Med, 1997. 30(2): p. 163–166.

12. Chen, B. and L. Eichenfield, Pediatric anesthesia in dermatologic surgery: when hand-holding is not enough. Dermatol Surg, 2001. 27(12): p. 1010–1018.

13. Chowdhary, S., et al., Risk of topical anesthetic-induced methemoglobinemia: a 10-year retrospective case-control study. JAMA Intern Med, 2013. 173(9): p. 771–6.

14. Langley, G.L., et al., The Improvement Guide: A Practical Approach to Enhancing Organizational Performance. 2nd ed. 2009. San Francisco: Jossey-Bass Publishers.

15. Sherman, J., et al., Let Us Use LET: A Quality Improvement Initiative. Pediatr Emerg Care, 2016. 32(7): p. 440–3.

16. Cregin, R., et al., Improving pain management for pediatric patients undergoing nonurgent painful procedures. Am J Health Syst Pharm, 2008. 65(8): p. 723–7.

17. Pitetti, R.D., S. Singh, and M.C. Pierce, Safe and efficacious use of procedural sedation and analgesia by nonanesthesiologists in a pediatric emergency department. Arch Pediatr Adolesc Med, 2003. 157(11): p. 1090–6.

18. Lawrence, L.M. and S.W. Wright, Sedation of pediatric patients for minor laceration repair: Effect on length of emergency department stay and patient charges. Pediatr Emerg Care, 1998. 14(6): p. 393–5.

19. Mellion, S.A., et al., Evaluating Clinical Effectiveness and Pharmacokinetic Profile of Atomized Intranasal Midazolam in Children Undergoing Laceration Repair. J Emerg Med, 2017. 53(3): p. 397–404.

